# Efficacy of *Lactococcus lactis* strain plasma (LC-Plasma) in easing symptoms in patients with mild coronavirus disease 2019 (COVID-19): protocol for an exploratory, multicenter, double-blinded, randomized controlled trial (PLATEAU study)

**DOI:** 10.1101/2022.02.13.22270913

**Authors:** Kazuko Yamamoto, Naoki Hosogaya, Tsuyoshi Inoue, Kenta Jounai, Ryohei Tsuji, Daisuke Fujiwara, Katsunori Yanagihara, Koichi Izumikawa, Hiroshi Mukae

## Abstract

**Introduction:** The coronavirus disease 2019 (COVID-19) pandemic has been a major concern worldwide; however, easily accessible treatment options for patients with mild COVID-19 remains limited. Since oral intake of *Lactococcus lactis* strain Plasma (LC-Plasma) enhances both the innate and acquired immune systems through activation of plasmacytoid dendritic cells (pDCs), we hypothesized that the oral intake of LC-Plasma could aid the relief or prevention of symptoms in patients with asymptomatic or mild COVID-19.

**Methods and analysis:** This is an exploratory, multicenter, double-blind, randomized, placebo-controlled trial. This study was initiated in December 2021 and concludes in April 2023. The planned number of enrolled subjects is 100 (50 patients × 2 groups); subject enrolment will be conducted until October 2022. Patients with asymptomatic or mild COVID-19 will be enrolled and randomly assigned in a 1:1 ratio to Group A (oral intake of LC-Plasma-containing capsule, 200 mg/day, for 14 days) or Group B (oral intake of placebo capsule, for 14 days). The primary endpoint is the change in subjective symptoms measured by the severity score. Secondary endpoints include SARS-CoV-2 viral loads, biomarkers for pDC activation, serum SARS-CoV-2-specific antibodies, serum cytokines, interferon and interferon-inducible antiviral effectors, and the proportion of subjects with emergency room visits to medical institutions or who are hospitalized.

**Ethics and dissemination:** The study protocol was approved by the Clinical Research Review Board of Nagasaki University, in accordance with the Clinical Trials Act of Japan. The study will be conducted in accordance with the Declaration of Helsinki, Clinical Trials Act, and other current legal regulations in Japan. Written informed consent will be obtained from all participants. The results of this study will be reported in journal publications.

**Registration:** This study was registered in the Japan Registry of Clinical Trials (registration number: jRCTs071210097).

**ARTICLE SUMMARY:** *Strengths and limitations of this study:*

- This is the first randomized controlled trial to assess the efficacy of *Lactococcus lactis* strain Plasma (LC-Plasma) in preventing the onset and attenuation of symptoms in patients with asymptomatic or mild COVID-19.
- This study is also the first to evaluate the significance of pDC-related immune responses, including interferon production, clearance of symptoms, and prevention of COVID-19 progression.
- The results of this study may contribute to the development of novel treatment options for asymptomatic or mild COVID-19 patients.
- This is an exploratory study, due to the lack of previous clinical evidence that evaluated the effect of LC-Plasma intake in patients with COVID-19.
- Other limitations include the subjective endpoint as the primary endpoint and generalizability, since this study will be conducted only in Japan in Japanese patients.

## INTRODUCTION

The coronavirus disease 2019 (COVID-19) pandemic is a major concern worldwide. In Japan, a cumulative total of 2.92 million polymerase chain reaction-positive cases have been confirmed, and 18,387 deaths have been reported by the Ministry of Health, Labor and Welfare in Japan as of December 28, 2021.[1] Although the extensive introduction of SARS-CoV-2 vaccines has effectively decreased the frequency of COVID-19-related severe diseases, breakthrough infections due to new SARS-CoV-2 variants are currently emerging threats.[2, 3]

Approximately 90% of COVID-19 patients have mild disease and symptoms in Japan.[4] From a public health perspective, the management of asymptomatic or mild cases is particularly important because these patients move around freely and may be unaware that they are infected with SARS-CoV-2, causing the spread of infection. Recently, monoclonal antibody therapies, such as casirivimab/imdevimab antibody cocktails, and novel oral antiviral agents, such as molnupiravir, have been approved for mild-to-moderate COVID-19.[5–7] These treatments are associated with reduced risk of hospitalization or death in patients with mild-to-moderate COVID-19.[8, 9] However, these new drugs are not easy to access and are restricted by the government for limited use in COVID-19 patients with high-risk backgrounds.

*Lactococcus lactis* strain plasma (LC-Plasma; e.g., *Lactococcus lactis* subsp. *lactis* JCM 5805) is a lactic acid bacterium that directly activates plasmacytoid dendritic cells (pDCs) and induces type I and III interferons (IFNs) through toll-like receptor 9 (TLR9) stimulation.[10, 11] pDCs are known to act as key regulators of antiviral immunity.[12] Activation of pDCs induces two antiviral immune responses: 1) production of IFN-alpha/beta to directly inhibit viral replication as part of the innate immune response [13] and 2) subsequent activation of adaptive T cell/B cell-mediated acquired immune response. [14] IFN production is a critical immune response that occurs rapidly during the early phase of SARS-CoV-2 infection.[15] One reason why elderly patients with COVID-19 have more severe disease and fatality than younger adults is insufficient IFN production in the early phase of infection. Early IFN production by the host accelerates the decrease in SARS-CoV-2 viral load and suppresses progression to severe disease.[16] This suggests that a novel treatment that activates innate immunity may help to prevent or suppress the onset of COVID-19 symptoms. A previous study reported that healthy volunteers who ingested LC-Plasma showed activated pDCs with increased activity of IFNs by peripheral blood mononuclear cells upon exposure to inactivated influenza virus and that onset of flu-like symptoms was prevented.[13] LC-Plasma treatment also increased the proportion of antigen-specific CD8^+^ T cells[14] whose coordination with CD4^+^ T cells is important for preventing the exacerbation of COVID-19.[17] These findings suggest that intake of LC-Plasma prevents and/or attenuates COVID-19 symptoms through the activation of both innate and acquired immune responses. Further, LC-Plasma has the advantage of being a natural and safe product according to previous reports.[18, 19]

Thus, LC-Plasma is a safe and easy-to-use candidate for treating asymptomatic or mild COVID-19. The present study aimed to evaluate the efficacy of LC-Plasma capsules in preventing the onset and attenuating symptoms of COVID-19 in patients with asymptomatic or mild infection.

## METHODS AND ANALYSIS

### Study design and setting

The “efficacy of *Lactococcus lactis* strain <u>PL</u>asmA <u>T</u>o <u>EA</u>se symptoms in patients with coronavir<u>U</u>s disease 2019 (PLATEAU) study: a multicenter, double-blinded, randomized placebo-controlled trial” was initiated in December 2021 following approval by the Clinical Research Review Board of Nagasaki University in November, 2021 (approval number: CRB20-027). This study was registered with the Japan Registry of Clinical Trials (registration number: jRCTs071210097) in December 2021 prior to study initiation. Patients have been enrolled since December 2021; enrolment will culminate in October 2022, and the study is scheduled to end in April 2023. This study is being conducted at seven medical institutions in Japan: Nagasaki University Hospital, Nagasaki Harbor Medical Center, Japanese Red Cross Nagasaki Genbaku Hospital, Saiseikai Nagasaki Hospital, Juko Memorial Nagasaki Hospital, Kouseikai Hospital, and Nagasakikita Tokushukai Hospital. As shown in Figure 1, eligible patients will be asked to participate in this study, and informed consent will be obtained prior to registration/randomization. After written consent is obtained from the eligible patients, they will be enrolled and randomized into Group A (intake of LC-Plasma-containing capsule) or Group B (intake of placebo capsule). Group A patients will be treated with 200 mg/day of heat-killed LC-Plasma, which includes at least 4.0 × 10^11^ cells.

**Figure 1.**
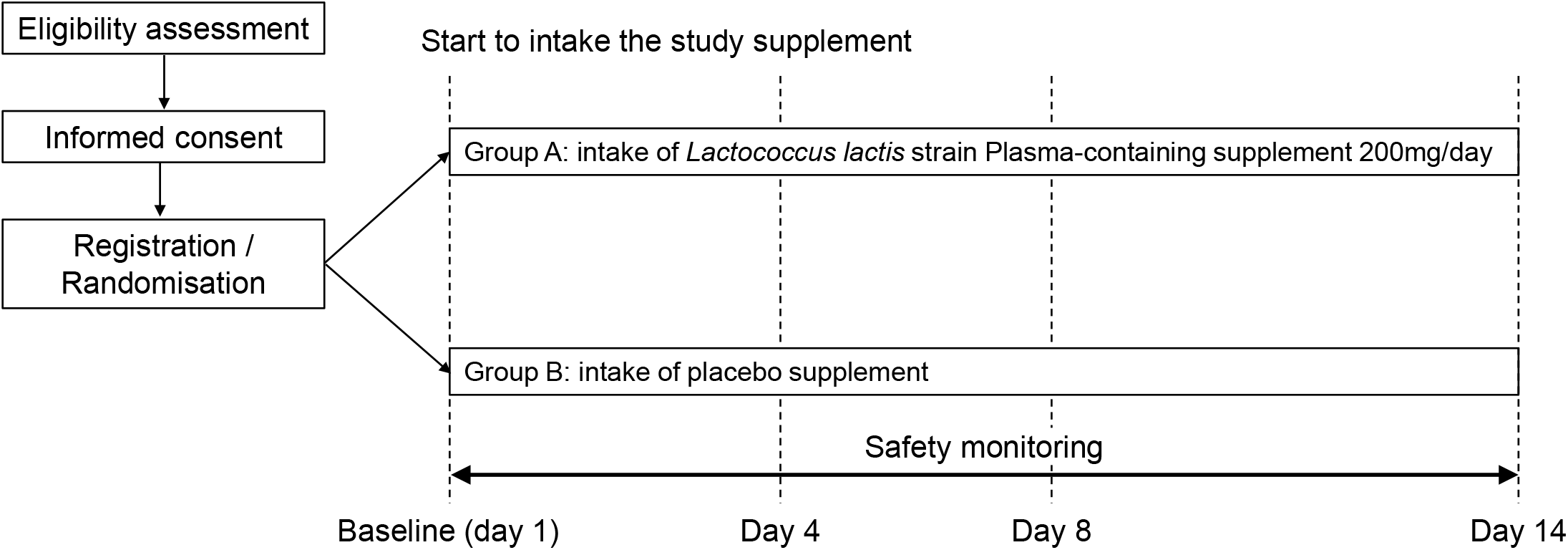
Study design and flow of recruitment, randomization, study intervention, and observation.

### Formulation of test capsules

A LC-Plasma-containing capsule contained 50 mg heat-killed LC-Plasma, 127.3 mg dextrin, and 2.7 mg calcium stearate. A placebo capsule contained 177.3 mg dextrin and 2.7 mg calcium stearate. Subjects will orally ingest 4 test capsules (LC-Plasma-containing capsules or placebo capsules) once daily. The study controller has confirmed that the two types of test capsules cannot be distinguished on the basis of taste, appearance, or smell.

### Eligibility criteria

Patients who are SARS-CoV-2 positive with asymptomatic or mild COVID-19 disease and who are stayed at isolation facilities for COVID-19 patients in Nagasaki City, Nagasaki, Japan are eligible for this study. The detailed inclusion criteria are as follows: 1) subjects who are SARS-CoV-2 positive, 2) whose arterial oxygen saturation (SpO_2_) is 96% or higher, 3) who are aged 20 years or older and younger than 65 years, 4) who do not refuse to disclose their vaccination status, 5) who can be stayed at isolation facilities for COVID-19 patients designated by Nagasaki City, and 6) who give their written consent to participate in the study. Subjects who meet any of the following exclusion criteria will be excluded from participation: 1) obese (body mass index (BMI) ≥30 kg/m^2^), 2) subjects with strong dyspnea, chest pain, or hemosputum, 3) previous history of COVID-19, 4) being treated or planning to be treated with neutralizing antibody drugs for SARS-CoV-2, 5) being treated with immunosuppressive agents, antirheumatic agents, corticosteroids, or immunoglobulin preparations, 6) subjects administered oral intestinal regulators, 7) taking one or more beverage or food containing LC-Plasma or yogurt that contains *Lactobacillus delbrueckii* subsp. *bulgaricus* daily, 8) who are pregnant, possibly pregnant, or breastfeeding, 9) participating in other clinical trials, 10) requiring legal representation for giving consent, or 11) with other conditions that the responsible investigator or sub-investigators deem inappropriate for study participation.

### Recruitment and consent

The informed consent document (see supplementary material) will be provided to candidates who meet all inclusion criteria and none of the exclusion criteria to provide a comprehensive explanation of this study. Written consent will be obtained from all participants. After obtaining consent, candidates will be provisionally enrolled in this study. Additional candidates will be enrolled in this study when they enter isolation facilities as described above.

### Random allocation

After obtaining informed consent, eligible participants will be randomly assigned in a 1:1 ratio to Group A (intake of LC-Plasma-containing capsule, 200 mg/day) or Group B (intake of placebo capsule). The randomization sequence will be generated using a computer-based dynamic allocation method with a minimization procedure to balance allocation factors (age: less than 50 years or 50 years or older; SARS-CoV-2 vaccination status; use of anti-SARS-CoV-2 agents).

### Blinding and anonymization

This study will be conducted as a double-blinded trial. All parties are blinded, including investigators, participants, the manufacturer of test capsules, core laboratories, and biostatisticians. A central registration number will be used to identify subjects for anonymization. The manufacturer of test capsules marked LC-Plasma-containing capsules and placebo capsules with specific characters such as X and Y, and the person in charge of assignment will complete a corresponding table of characters and central registration numbers. The computer-based allocation system will provide a central registration number linked to each subject during registration/assignment. Unblinding will be conducted only in emergency cases such as the occurrence of serious adverse events (SAEs).

### Study intervention and observation

Subjects will be assigned to a group and orally administered test capsules for 14 days. All subjects will be subsequently observed for 14 days (observation points are days 1, 4, 8, and 14). During the observation period, subjects will be restricted to using systemic antiviral agents (except topical antiviral agents) or intestinal regulators. For palliative care for COVID-19, temporary use of antitussive agents, expectorants, general cold medications, antipyretic analgesic (acetaminophen only), and antidiarrheal agents are allowed, but not for routine use. In addition, during the observation period, addition, discontinuation, switch, or dose change of any medical agents or supplements will be recorded. Furthermore, subjects will be asked to refrain from consuming lactic acid bacteria-containing foods, such as yogurt, during the observation period. The intervention will be discontinued if the subjects meet any of the discontinuation criteria: consent withdrawal, worsening of the primary disease, complications, adverse events (AEs) requiring discontinuation of the study intervention, remarkably poor adherence to the test capsules regimen, or any other condition that investigators deem appropriate for discontinuation of the study intervention.

Table 1 shows the schedule of assessments to be performed at each observation point, including mandatory and optional assessments. The observation items are as follows: 1) subjects’ background characteristics (sex, height, weight, BMI, presence or absence of onset of COVID-19, estimated infection day, onset day of COVID-19 if symptomatic, SARS-CoV-2 vaccination status (timing, number of vaccinations, vaccine type, last day of vaccination), 2) Severity of COVID-19 assessed by severity classification according to COVID-19 infectious disease treatment guidelines published by the Ministry of Health, Labor and Welfare in Japan [20] (mild: SpO_2_ ≥ 96%, no respiratory symptoms or only coughing without dyspnea, and without pneumonia findings, moderate I: 93% < SpO_2_ < 96%, with dyspnea, and with pneumonia findings, moderate II: SpO_2_ ≤ 93% and oxygen administration required, and severe: admission to intensive care unit or ventilator required), 3) Blood Test 1 (differential count of leukocytes (neutrophil, lymphocyte, eosinophil, monocyte, and basophil), platelet, aspartate aminotransferase, alanine aminotransferase, gamma-glutamyl transpeptidase, blood urea nitrogen, serum creatinine, lactate dehydrogenase, C-reactive protein, ferritin, and HbA1c), 4) Blood Test 2 (pDCs activation markers (human leukocyte antigen DR and CD86), cytokines (interleukin-6 and monocyte chemoattractant protein-1), SARS-CoV-2 specific antibodies (immunoglobulin M and G), expression of interferon and interferon-inducible antiviral effector genes in PBMC, and immune cell analysis (T cells and B cells), 4) real-time polymerase chain reaction test for SARS-CoV-2 by nasopharyngeal swabs, 5) medication information, 6) vital signs (body temperature, pulse, SpO_2_, and frequency of breath), 7) adherence of test capsules, 8) patients’ subjective symptoms measured by the severity score [21] and visual analog scale (VAS), and 9) AEs/SAEs.

**Table 1.**
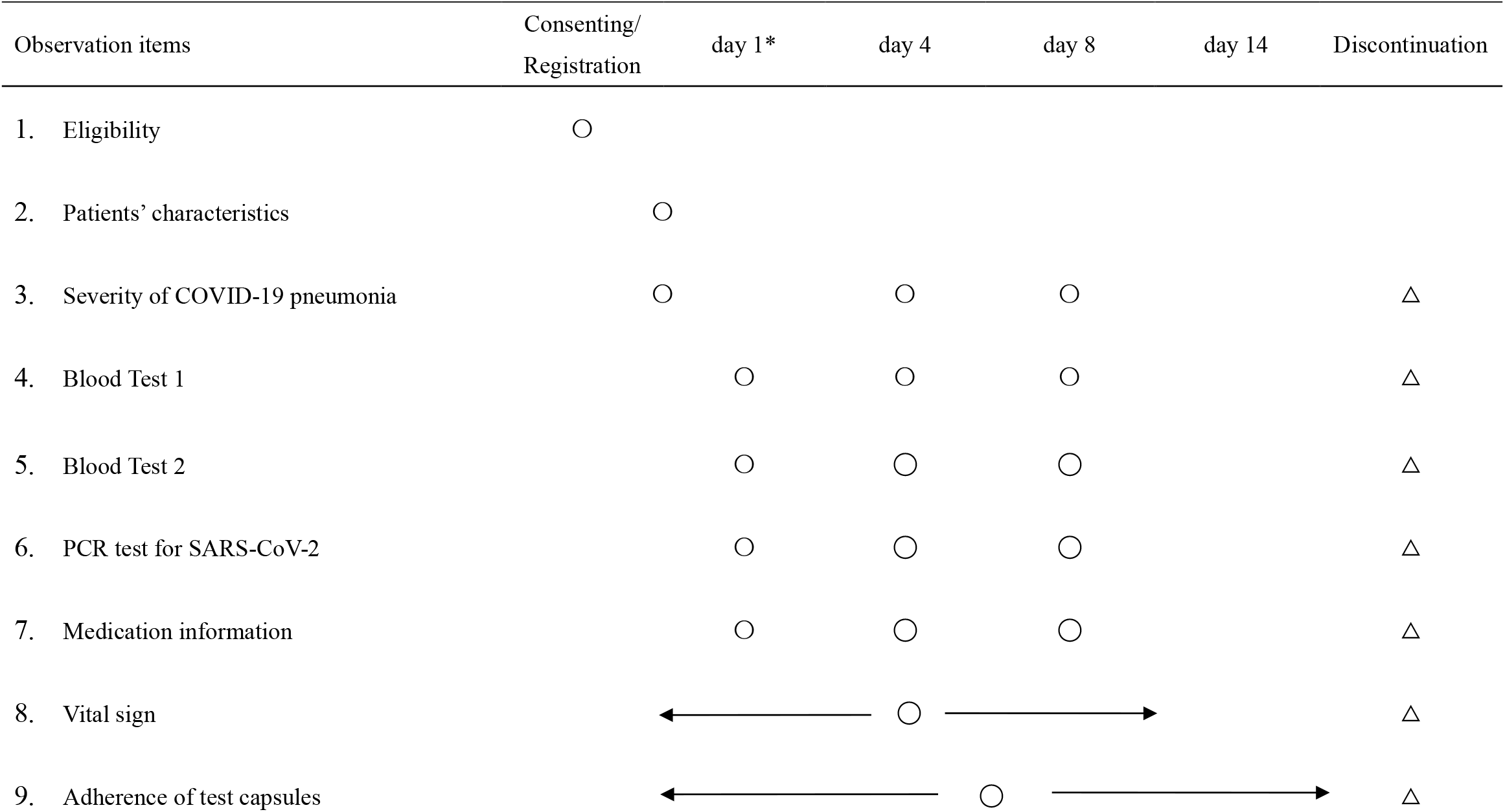
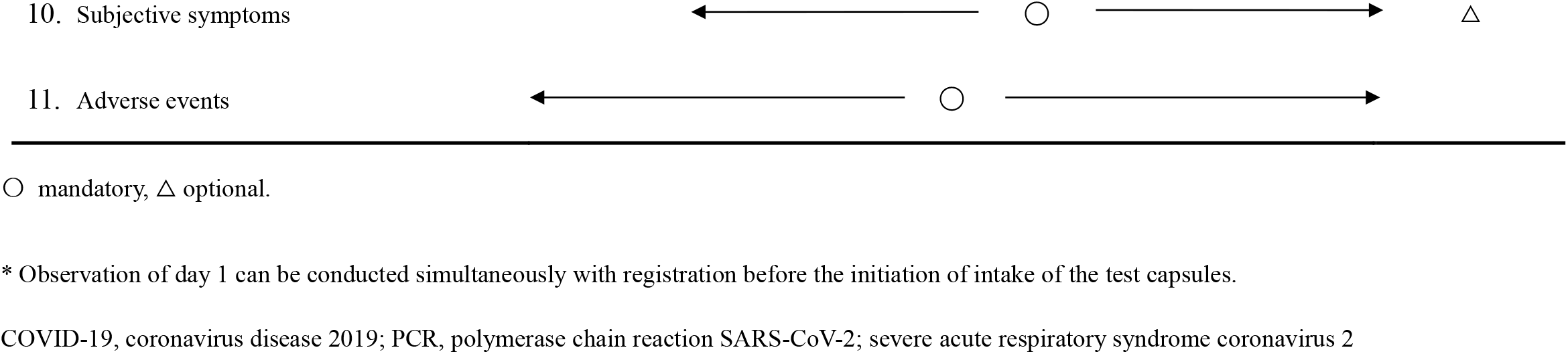
Schedule of assessments performed at each observation point

### Outcomes

The primary endpoint of this study is the change in subjective symptoms measured using the severity score [21] and Visual Analogue Scale (VAS). Subjective symptoms (cough, shortness of breath, fatigue, headaches, anosmia, dysgeusia, and anorexia) are assessed using a 4-point Likert scale (not affected = 0 points, little effect = 1 point, affected = 2 points, and severely affected = 3 points), and the total severity score is calculated as the sum of scores for these seven subjective symptoms. The severity score questionnaire is shown in Supplementary Table 1.

Secondary endpoints are as follows: 1) change or percent change in viral load of SARS-CoV-2, 2) change or percent change in biomarkers for pDC activation (HLA-DR, CD86), 3) change or percent change in SARS-CoV-2-specific antibodies (immunoglobulin G and M), 4) change or percent change in blood cytokines (IL-6, MCP-1), 5) change or percent change in IFN-or IFN-inducible antiviral effectors, and 6) proportion of subjects who visit the emergency room and are hospitalized.

### Data collection, data management, and monitoring

A case report form will be used for data collection. Data collection and management will be carried out by third party entities to avoid bias. Data will be managed by Soiken Inc., the Data Management Group (Data Center). To ensure quality, the study will be monitored by Soiken Inc., the Monitoring Group.

### Safety evaluation

During this study, investigators will constantly monitor for any AEs during regular medical checkups. All related AEs, including study agent side effects, abnormal clinical laboratory test values, and untoward medical occurrences, will be reported and documented. If AEs meet the following criteria, they are referred to as SAEs based on the ICH E2A, ICH E2D, and “Ethical Guidelines for Medical and Health Research Involving Human Subjects”:[22] AEs that result in 1) death, 2) life-threatening hospitalization, or 3) extension of hospitalization, 4) persistent or significant disability or incapability, 5) medically important or critical condition, 6) AEs that are equivalently severe to criteria to 5), or 7) AEs that cause congenital abnormality or birth defects. AEs will be followed until normalization or recovery to a level that is not considered an AE.

### Sample size calculation

The severity score was used in a case series study to estimate the patients’ subjective symptoms.[21] When the total severity score from this previous study was translated using a calculation method mentioned in the outcomes section of the present study, the total score on days 0 and 14 was 11 ± 5.3 and 1.7 ± 2.3, respectively, and the change from day 0 to day 14 was –9.3 ± 6.0.

Since this study will enroll patients who are asymptomatic or have mild COVID-19, we assumed that the severity score of cough at baseline (day 1) will be 0 to 1 and that of other symptoms will be 0 in this study. Therefore, the mean total severity score at baseline predicted in this study is 0.5. We also assumed that the severity score will not worsen in Group A (i.e., change in the total severity score = 0). Since a recent meta-analysis reported that 48.9% of initially asymptomatic patients with COVID-19 became symptomatic,[23] we assumed that the severity score in Group B will worsen to a level of half of the severity score at day 0 reported in the case series study mentioned above,[21] meaning that the total severity score would be 5.5 (i.e., change in the total severity score = 5). Since it is expected that the degree of subjective symptoms varies widely among patients when asymptomatic subjects become symptomatic, we assumed that the standard deviation in the change in the severity score will be 7.0, which is slightly larger than that in the case series study mentioned above.[21] Under these assumptions, the minimum sample size required to achieve a significance of 0.05 from a two-sided test with a statistical power of 90% was determined to be 42 subjects for both groups, or a total of 84 subjects. We estimated the dropout rate to be 15%; thus, planned enrolment was set at 100 subjects, with 50 in each group.

### Statistical analysis

All tests will be two-sided, and a *p*-value <0.05 will be considered statistically significant. As this is an exploratory trial, multiplicity will not be adjusted for all endpoints. A statistical analysis plan was developed prior to the database lock. All statistical analyses will be conducted by independent biostatisticians.

Three analysis sets are defined in this study; the full analysis set (FAS) includes all patients who will be registered in this study. However, patients with severe protocol violations, such as registration without obtaining consent or registration outside of the enrolment period, will be excluded from the FAS. The per-protocol set excludes patients with a protocol violation, such as violation of eligibility criteria, use of prohibited or restricted concomitant treatments, or poor adherence to the test capsules (less than 75% or more than 120%). The safety analysis set includes all patients who will be registered in this study who receive at least one dose of the test capsules.

Patient characteristics at baseline will be presented as frequencies and proportions for categorical data and summary statistics (number of patients, mean, standard deviation, minimum, first quartile, median, third quartile, and maximum) for continuous data. Patient characteristics will be compared using the chi-square test or Fisher’s exact test for categorical data and the two-sample t-test or Wilcoxon rank sum test for continuous variables.

The primary endpoint of this study is the change in subjective symptoms measured by the severity score.[21] The total score of the severity score is calculated by summarizing the severity scores of seven symptoms, and the summary statistics of change in the total score of the severity score from baseline to day 14 will be calculated. Analysis of covariance (ANCOVA) will be conducted to test the null hypothesis that the change in the total score of the severity score from baseline to day 14 is the same in both groups. The allocation factors age (less than 50 years or 50 years or older), vaccination status for SARS-CoV-2 (vaccinated or unvaccinated), use of anti-SARS-CoV-2 agent (presence or absence of anti-SARS-CoV-2 agent use), and the total score of the severity score at baseline will be used as covariates in the ANCOVA. The sensibility analysis will be performed using the mixed-effects model for repeated measures with an unstructured covariance structure with treatment groups, time (day), interactions between treatment groups and time, allocation factors, and the total score of the severity score at baseline as fixed effects, and subjects as random effects. If the calculation results do not converge, compound symmetry will be used. The VAS score will be analyzed similar to the analysis of severity score.

For the secondary endpoints, summary statistics (number of patients, mean, standard deviation, minimum, first quartile, median, third quartile, and maximum) for measurements, changes from baseline, and percentage changes from baseline will be calculated for continuous data. Frequencies and proportions will be calculated for categorical data. Two-sample *t*-test or Wilcoxon rank sum test for intergroup comparisons of continuous data, one-sample *t*-test or Wilcoxon signed-rank test for intragroup comparisons of continuous data, and the chi-square test or Fisher’s exact test for intergroup comparisons of categorical data will be performed.

For safety endpoints, summary statistics for the frequency of AEs will be calculated for each group, and Fisher’s exact tests will be performed for intergroup comparisons.

### Patient and public involvement

Patients and the public were not involved in the conception or planning of this study and will not be involved in the execution, analysis, and evaluation.

## DISCUSSION

This is the first randomized controlled trial to estimate the efficacy of LC-Plasma in preventing the onset and attenuating the symptoms of patients with asymptomatic or mild COVID-19. Since the intake of LC-Plasma enhances both the innate [13] and acquired immune responses [14] through the activation of pDCs,[10, 11] we hypothesize that the intake of LC-Plasma contributes to the prevention of the onset and attenuation of the severity of COVID-19. Easy-access treatment options for patients with mild COVID-19 are limited, thus, results of this study may contribute to the treatment of asymptomatic or mild COVID-19 patients. This study is also noteworthy because it evaluates the significance of pDC-related immune responses, including IFN production, clearance of symptoms, and prevention of COVID-19 progression.

The safety of LC-Plasma has been previously confirmed. Long-term intake tests (50 mg daily, 12 weeks, or 150 mg daily, 4 weeks) [18] and an excessive intake test (250 mg daily, 4 weeks) [19] of LC-Plasma reported no safety concerns. In addition, LC-Plasma-containing yogurt, beverages, and supplements have been commercially available since 2012, and no health hazards associated with LC-Plasma-containing products have been reported. This suggests the safety of the LC-Plasma-containing supplements used in this study (200 mg daily for 14 days). Molnupiravir, the first approved oral antiviral agent for high-risk COVID-19 patients, has common side effects including diarrhea, nausea, dizziness, and headache within 14 days of the last dose, and is restricted in pregnant women because it affects the development of the fetus [24, 25]. Non-elderly and non-high-risk patients with mild COVID-19 rarely progress to severe disease; therefore, it is reasonable to choose safer drugs that sufficiently control their symptoms.

This study has several limitations. First, this is an exploratory study due to the lack of previous clinical evaluation of the effects of LC-Plasma intake in patients with COVID-19. The target number of enrolled patients in this study was calculated from results of a case series study estimating subjective symptoms in non-hospitalized patients with COVID-19 treated with the histamine-2 receptor antagonist famotidine.[21] Since this case series evaluated subjective symptoms in only 10 patients, the calculated mean and standard deviation of the results might not reflect the actual patients’ subjective symptoms. Second, the primary endpoint in this study is patients’ subjective symptoms as reported by subjects themselves. Therefore, biases such as responder bias, non-responder bias, and volunteer bias cannot be completely avoided. However, the effect of bias is minimized using a double-blinded study design. Third, this study will be conducted in medical institutions in Japan and will enroll only Japanese patients. These constraints could limit the generalizability of this study. Further larger-scale, international clinical trials are required in the future.

### Ethics and dissemination

This study and its protocol were approved by the Clinical Research Review Board of Nagasaki University (Approval No. CRB20-027) in accordance with the Clinical Trials Act of Japan. The study will be conducted in accordance with the Declaration of Helsinki, Clinical Trials Act, and other current legal regulations in Japan. If any amendments on the protocol are required, the amended protocol will be resubmitted for investigation and approval by the Clinical Research Review Board of Nagasaki University prior to the implementation of this study according to the amended protocol. Written informed consent will be obtained from all participants after a full explanation of the study. Any health hazards caused by this study will be compensated by clinical research insurance. The results of this study will be disseminated through medical conferences and journal publications.

Datasets generated and/or analyzed during this study will not be publicly available because of the absence of a statement in the study protocol and the informed consent documents enabling data sharing with a third party after the end of the study. Data sharing has not been approved by the certified review board.

## Acknowledgements

The study is funded by Kirin Holdings Co., Ltd. The authors thank all clinical staff for their assistance in the execution of the study and Soiken Inc. for their technical assistance in the launch and execution of the study.

## Author contributions

Kazuko Yamamoto, Ryohei Tsuji, Kenta Jounai, and Daisuke Fujiwara contributed to the study conception and design, drafted the protocol, and supervised the revision. Naoki Hosogaya, Katsunori Yanagihara, Tsuyoshi Inoue, and Koichi Izumikawa provided intellectual input to improve the study design and to revise the protocol. Hiroshi Mukae supervised the conception and design of the study. All authors have read and approved the final manuscript.

## References

1. Ministry of Health, Labour and Welfare in Japan. Novel Coronavirus (COVID-19). Available at https://www.mhlw.go.jp/stf/seisakunitsuite/bunya/0000164708_00079.html. Last accessed at December 14, 2021

2. World Health Organization. WHO Coronavirus (COVID-19) Dashboard. Available at https://covid19.who.int/. Last accessed at December 14, 2021.

3. World Health Organization. Classification of Omicron (B.1.1.529): SARS-CoV-2 Variant of Concern. Available at https://www.who.int/news/item/26-11-2021-classification-of-omicron-(b.1.1.529)-sars-cov-2-variant-of-concern. Last accessed at December 14, 2021

4. Ministry of Health, Labour and Welfare in Japan. 11 things you need to know NOW about COVID-19 (December, 2021 version). Available at https://www.mhlw.go.jp/stf/seisakunitsuite/bunya/0000164708_00079.html. Last accessed at January 7th, 2022

5. Weinreich DM, Sivapalasingam S, Norton T et al. REGN-COV2, a Neutralizing Antibody Cocktail, in Outpatients with Covid-19. N Engl J Med 2021; 384: 238–251

6. Piccicacco N, Zeitler K, Montero J et al. Effectiveness of Severe Acute Respiratory Syndrome Coronavirus 2 Monoclonal Antibody Infusions in High-Risk Outpatients. Open Forum Infect Dis 2021; 8: ofab292

7. Koehler J, Ritzer B, Weidlich S et al. Use of monoclonal antibody therapy for nosocomial SARS-CoV-2 infection in patients at high risk for severe COVID-19: experience from a tertiary-care hospital in Germany. Infection 2021; 49: 1313–1318

8. Lee CC, Hsieh CC, Ko WC. Molnupiravir-A Novel Oral Anti-SARS-CoV-2 Agent. Antibiotics (Basel) 2021; 10: 1294

9. Medicines and Healthcare products Regulatory Agency in United Kingdom. First oral antiviral for COVID-19, Lagevrio (molnupiravir), approved by MHRA. Available at https://www.gov.uk/government/news/first-oral-antiviral-for-covid-19-lagevrio-molnupiravir-approved-by-mhra. Last accessed at December 14, 2021

10. Jounai K, Ikado K, Sugimura T et al. Spherical lactic acid bacteria activate plasmacytoid dendritic cells immunomodulatory function via TLR9-dependent crosstalk with myeloid dendritic cells. PLoS One 2012; 7: e32588

11. Fujiwata D. lactic acid bacterium that activates plasmacytoid dendritic cells. Experimental Medicine 2017; 35: 191–196

12. Kadowaki N, Antonenko S, Lau JY et al. Natural interferon alpha/beta-producing cells link innate and adaptive immunity. J Exp Med 2000; 192: 219–226

13. Sugimura T, Takahashi H, Jounai K et al. Effects of oral intake of plasmacytoid dendritic cells-stimulative lactic acid bacterial strain on pathogenesis of influenza-like illness and immunological response to influenza virus. Br J Nutr 2015; 114: 727–733

14. Suzuki H, Jounai K, Ohshio K et al. Administration of plasmacytoid dendritic cell-stimulative lactic acid bacteria enhances antigen-specific immune responses. Biochem Biophys Res Commun 2018; 503: 1315–1321

15. Shah VK, Firmal P, Alam A et al. Overview of Immune Response During SARS-CoV-2 Infection: Lessons From the Past. Front Immunol 2020; 11: 1949

16. Park A, Iwasaki A. Type I and Type III Interferons - Induction, Signaling, Evasion, and Application to Combat COVID-19. Cell Host Microbe 2020; 27: 870–878

17. Rydyznski Moderbacher C, Ramirez SI, Dan JM et al. Antigen-Specific Adaptive Immunity to SARS-CoV-2 in Acute COVID-19 and Associations with Age and Disease Severity. Cell 2020; 183: 996–1012.e19

18. Tanaka K, Suzuki A, Kanayama M et al. The safety evaluation of long-term or excessive intake of the beverage containing Lactococcus lactis subsp. lactis JCM 5805 and Resistant Maltodextrin - A randomized, double-blind, placebo-controlled, parallel-group trial-. Jpn Pharmacol Ther. 2015; 43: 1711–1727

19. Kato Y, Kanayama M, Yanai S et al. Safety Evaluation of Excessive Intake of Lactococcus lactis Subsp. lactis JCM 5805: A Randomized, Double-Blind, Placebo-Controlled, Parallel-Group Trial. Food Nutr Sci 2018; 9: 403

20. Ministry of Health, Labour and Welfare in Japan. COVID-19 infectious disease treatment guidelines (ver.5.1). Available at https://www.mhlw.go.jp/stf/seisakunitsuite/bunya/0000121431_00111.html. Last accessed at August 2nd, 2021. (in Japanese)

21. Janowitz T, Gablenz E, Pattinson D et al. Famotidine use and quantitative symptom tracking for COVID-19 in non-hospitalised patients: a case series.. Gut 2020; 69: 1592–1597

22. Ministry of Health, Labour and Welfare in Japan. Ethical Guidelines for Medical and Health Research Involving Human Subjects. Available at https://www.mhlw.go.jp/stf/seisakunitsuite/bunya/hokabunya/kenkyujigyou/i-kenkyu/index.html. Last accessed at February 12th, 2021. (in Japanese)

23. He J, Guo Y, Mao R et al. Proportion of asymptomatic coronavirus disease 2019: A systematic review and meta-analysis. J Med Virol 2021; 93: 820–830

24. U.S. Food & Drug Administration. Important Safety Information Regarding Use of Molnupiravir in Pregnancy and Individuals of Childbearing Potential. Available at https://www.fda.gov/media/155101/download. Last accessed at January 7th, 2022

25. U.S. Food & Drug Administration. Coronavirus (COVID-19) Update: FDA Authorizes Additional Oral Antiviral for Treatment of COVID-19 in Certain Adults. Available at https://www.fda.gov/news-events/press-announcements/coronavirus-covid-19-update-fda-authorizes-additional-oral-antiviral-treatment-covid-19-certain. Last accessed at January 7th, 2022

