## Supplemental Table 1 for "Efficacy of *Lactococcus lactis* strain plasma (LC-Plasma) in easing symptoms in patients with mild coronavirus disease 2019 (COVID-19): protocol for an exploratory, multicenter, double-blinded, randomized controlled trial (PLATEAU study)"

Supplementary Table 1 Severity Score Questionnaire

<Subjective symptoms>

How are you affected by the following symptoms in your daily activities, compared with before the infection of the new coronavirus?

Select only one option for each symptom.

| | | |
|---|---|---|
| Subjective symptoms | Cough | □0: not affected, □1: little affected, □2: affected, □3: severely affected |
| | Shortness of breath | □0: not affected, □1: little affected, □2: affected, □3: severely affected |
| | Fatigue | □0: not affected, □1: little affected, □2: affected, □3: severely affected |
| | Headaches | □0: not affected, □1: little affected, □2: affected, □3: severely affected |
| | Anosmia | □0: not affected, □1: little affected, □2: affected, □3: severely affected |
| | dysgeusia | □0: not affected, □1: little affected, □2: affected, □3: severely affected |
| | Anorexia | □0: not affected, □1: little affected, □2: affected, □3: severely affected |
